# Rethinking wellbeing measurement: learning from a scale development study with adolescents living with HIV in Zimbabwe

**DOI:** 10.64898/2026.06.25.26356625

**Authors:** Webster Mavhu, Jermaine M. Dambi, Victoria Simms, Faith Martin, Joni Lariat, Rufaro Mbundure, Leviticus Makoni, Laura Kafata, Carol Wogrin, Vivian Chitiyo, Abigail Mutsinze, Frances M. Cowan, Nicola Willis, Sarah Bernays

**Author notes:** Joint first authors (Webster Mavhu, Jermaine M. Dambi).

## Abstract

Adolescents living with HIV (ALHIV) in Eastern and Southern Africa experience substantial challenges around medication adherence, treatment, care, mental health and wellbeing. Although interventions such as Zvandiri aim to strengthen mental health and wellbeing to impact HIV treatment outcomes, available measures often focus on negative outcomes or rely on positive psychology instruments developed in high-income settings. These tools may not adequately capture culturally embedded, relational and fluctuating dimensions of wellbeing among ALHIV. We describe the development of the Zvandiri Character Strength (ZCS) scale and draw methodological lessons for culturally grounded wellbeing measurement.

The multi-stage scale-development process (2023-2025) comprised a scoping and systematic review, qualitative interviews and focus group discussions, expert panel input, construct and item prioritisation with ALHIV, cognitive interviews, iterative item reduction, and exploratory factor and Rasch analyses. An initial item bank was reduced to a 53-item candidate scale, then to the ZCS 38 and finally ZCS 25, based on quantitative analyses and participant feedback. The ZCS 25 was subsequently assessed using retrospective pretest, post-test and follow-up administration.

The review identified conceptually overlapping positive psychological constructs, many of which are measured using instruments developed in high-income settings and not adequately validated for ALHIV in sub-Saharan Africa. Qualitative work highlighted connectedness, happiness, hope, motivation, optimism, and perseverance as key. In addition, discussions highlighted how wellbeing was relational, contextually situated, and often co-occurring with distress. Quantitative testing showed highly positive responses, disordered thresholds, ceiling effects, weak internal structure and fluctuating retrospective ratings. Participant feedback suggested that positive responses were likely due to affirmations and partly linked to social desirability.

In conclusion, the ZCS scale did not support a single interpretable total score, but the process of its development offers important methodological lessons. Future measurement should begin with clearer construct definition, response-process validation, domain-level testing, and explicit decisions about whether the measure aims to capture lived wellbeing, domain-specific positive outcomes or intervention-relevant change.

## Introduction

Globally, adolescents living with HIV (ALHIV) have the highest rates of attrition from HIV treatment and care of any age group, resulting in higher rates of treatment failure, morbidity and mortality compared to children and adults [1, 2]. In Eastern and Southern Africa, which is home to 84% of ALHIV globally, adolescents are the only population with increasing mortality rates [3, 4]. Elevated mortality rates are exacerbated by the significant prevalence of common mental health conditions [5]. Poor mental health is important in its own right, but also because it impacts adherence to HIV medication. For example, depression and anxiety significantly hinder treatment adherence, which in turn results in higher mortality [6].

Zvandiri (meaning ‘As I am’ in Shona, www.zvandiri.org) is a theoretically-grounded, multi-component differentiated service delivery model for children, adolescents and young adults living with HIV (CAYALHIV) [2, 7, 8]. Zvandiri links young people living with HIV to Community Adolescent Treatment Supporters (CATS); peer counsellors who are also living with HIV. These trained and mentored supporters help improve CAYALHIV’s health, wellbeing, and engagement with HIV treatment and care [2, 9–11]. In a cluster-randomised trial of the Zvandiri model, ALHIV receiving the Zvandiri intervention had 42% lower prevalence of virological failure or death after 2 years compared to ALHIV receiving standard care alone (95% CI: 0.36-0.94) [2, 12, 13]. A subsequent trial assessed the effect of training CATS in problem-solving therapy and found a five-fold reduction in the odds of common mental disorder symptoms [14].

Our past research suggests that improved mental health and wellbeing enable medication adherence [15, 16]. Nevertheless, a gap persists in the comprehensive evaluation of wellbeing, since most of the research to date prioritises negative indicators such as depression and anxiety [17, 18]. Of note, reliance on measuring negative outcomes undermines our capacity to identify and quantify the development of protective attributes, as proactive prevention.

Positive constructs have not been wholly absent from ALHIV research. Our review identified a range of positive psychological outcomes already used in this field, including resilience, self-esteem, self-efficacy, self-worth, coping, flourishing and positive outlook [17]. The problem is that this literature is conceptually fragmented, with overlapping constructs often measured with instruments developed in high-income settings that are not adequately adapted or validated for ALHIV in Eastern and Southern Africa [19]. The task, therefore, is to develop culturally appropriate, meaningful and psychometrically defensible approaches to assessing the positive outcomes and mechanisms through which interventions may improve young people’s lives.

ALHIV interventions may plausibly affect outcomes such as belonging, self-worth, treatment confidence, future orientation, resilience and relational safety, even where symptom-based or virological outcomes do not fully capture change. Measuring these domains is therefore important, but it requires clarity about which construct is being assessed, why it matters to the intervention theory of change, and whether the measure is valid for the language, context and social conditions in which young people are responding.

Measurement of wellbeing is shaped by three overlapping traditions: hedonic or subjective wellbeing, typically indexed through affect and life satisfaction; eudaimonic or psychological wellbeing, which emphasises positive functioning, growth, purpose and self-acceptance; and social wellbeing, which foregrounds social integration, contribution, coherence, actualisation and acceptance [20]. These traditions are related, but not interchangeable, and recent reviews confirm that overall wellbeing remains conceptually heterogeneous and is still measured using partly divergent frameworks, most of which were developed and validated in high-income settings [20–23]. High-income settings are more often characterised by individualistic cultures, which may place greater emphasis on eudaimonic than social wellbeing [17, 24].

Promoting positive mental health remains essential for improving ALHIV’s wellbeing and long-term outcomes [17, 25]. Intervention benefits may manifest as gains in belonging, recognition, future orientation, self-worth, or treatment confidence, even when distress remains. [17, 24]. We undertook a range of initiatives in collaboration with ALHIV to co-design a youth-focused, locally relevant quantitative positive psychology instrument; the Zvandiri Character Strength (ZCS) scale. The ZSC is a combination of learned and learnable attributes, virtues, skills, habits or capabilities that enable ALHIV to lead better lives.

Here, we report on our challenges and learnings to inform future initiatives. We frame the ZCS scale development process as a methodological case study within the psychometrics, patient-reported outcome measures, and wellbeing measurement literatures. The contribution is not to present relational wellbeing, response-process issues, social comparison, response shift, construct clarity or cross-cultural equivalence as new concerns; these are already well-established [26–30]. Rather, the ZCS case shows how these concerns became practically consequential when a relational HIV intervention in Zimbabwe required an interpretable positive wellbeing outcome measure.

## Methods

Fig 1 outlines the various steps of the ZCS scale development process.

**Fig 1:**
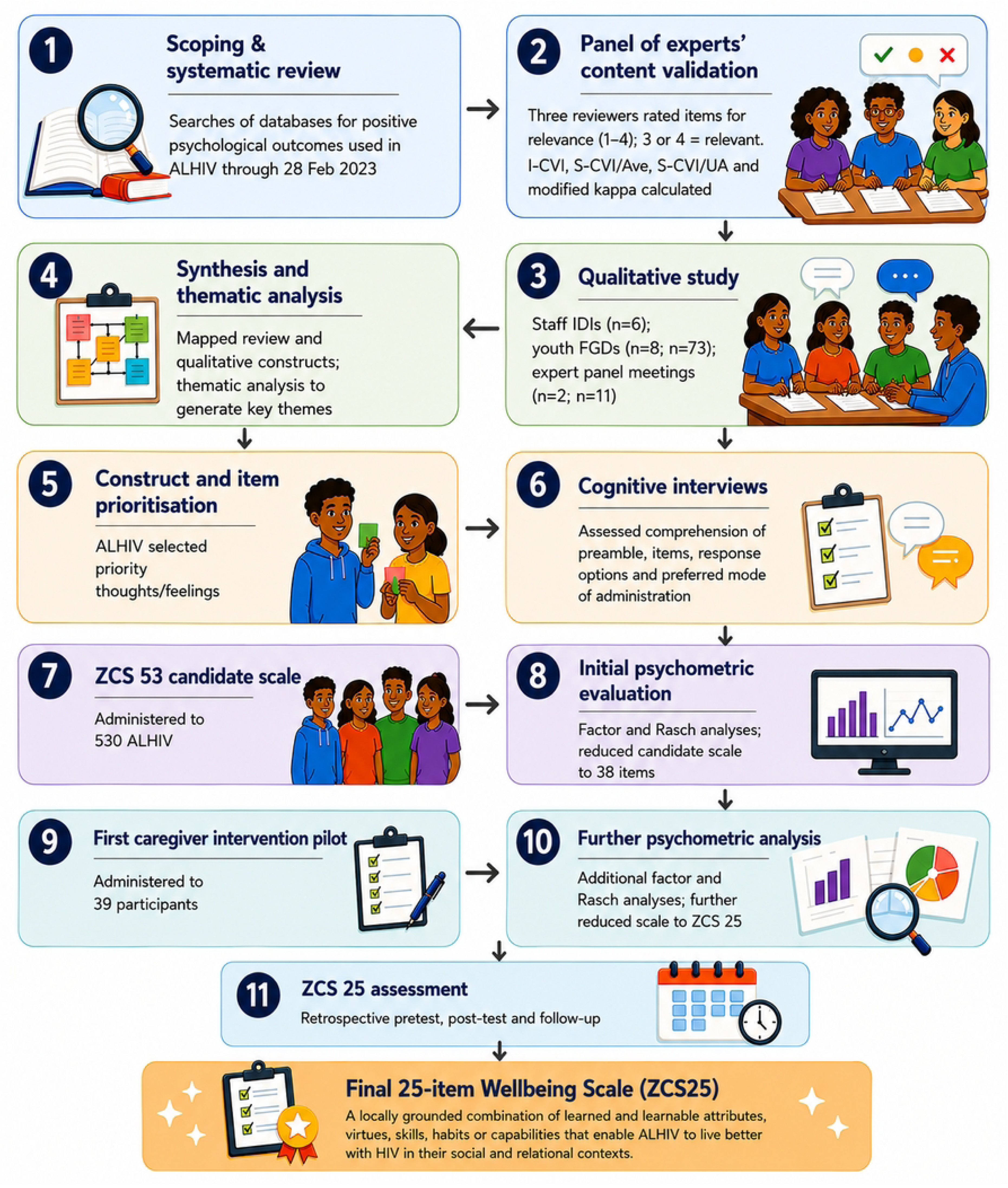
Steps leading to the 25-item wellbeing scale (ZCS 25)

We began by conducting a scoping and systematic review of positive psychological outcomes used in ALHIV (Fig 1, step 1) [17, 31]. In brief, we searched relevant databases for articles through 28 February 2023. We initially planned to only extract items from outcome measures with robust validity and reliability. However, only a few outcome measures had been properly adapted and validated for use in low- and middle-income countries [17]. Consequently, we adopted a pragmatic approach by selecting items from measures with apparent content validity and using thematic analysis to identify and group conceptually equivalent or duplicate items, resulting in an initial 263-item bank. Afterwards, three reviewers independently rated each item for relevance on a four-point scale, with ratings of 3 or 4 considered relevant. Content validity was assessed using the item-level content validity index (I-CVI) and scale-level indices (S-CVI/Ave and S-CVI/UA), with modified kappa used to adjust for chance agreement. Items with I-CVI = 1.00 were retained, those with I-CVI = 0.67 were revised, and those with I-CVI ≤ 0.33 were considered for removal; repeated or incomplete entries were excluded (Fig 1, step 2) [32].

We then carried out a complementary qualitative study that included six in-depth interviews with Zvandiri staff, eight focus group discussions with young people (n=73), and two expert panel meetings with peer supporters and beneficiaries (n=11) (Fig 1, step 3). We applied narrative synthesis to map the key constructs emerging from the scoping review and conducted interpretive thematic analysis to generate themes from the qualitative data (Fig 1, step 4). We then worked with two separate groups of ALHIV (n=24) to identify the constructs/items they felt conveyed the most important thoughts or feelings that Zvandiri should aim for to help them think and feel their best (Fig 1, step 5). We consolidated both groups’ rankings, captured discussions of their thought processes, and analysed them qualitatively. The final product was a 53-item scale spanning 14 constructs (S1 File). Items were scored on a Likert scale with responses ‘strongly disagree’, ‘disagree’, ‘agree’, and ‘strongly agree’.

Next, we conducted a series of cognitive interviews to assess comprehension of the scale’s preamble, whether scale items were understood as intended, whether the scale response options were working as intended, and the preferred mode of scale administration (Fig 1, step 6). We then administered the 53-item scale to 530 ALHIV using an interviewer-administered card-ranking process (Fig 1, step 7). The group consisted of 240 Zvandiri beneficiaries and 290 ALHIV who had not previously been in contact with Zvandiri and were accessing treatment at a Harare tertiary facility. We held a group discussion with young people (n=12) to investigate emerging issues.

We used both factor and Rasch analyses to reduce the scale to 38 items (Fig 1, step 8). Next, we administered the ZCS 38 scale to 39/40 young people who had been part of a pilot intervention exploring whether targeting caregivers improved a young person’s relational environment and, therefore, their wellbeing, to glean evidence of the scale’s feasibility, particularly regarding respondent burden (Fig 1, step 9). Following additional factor and Rasch analyses, we further reduced the items to 25, hence the ZCS 25 scale (Fig 1, step 10). Lastly, we administered the ZCS 25 scale to eight young people who had been part of a second pilot caregiver intervention phase, initially asking them to think back to before they began the programme (retrospective pretest) and rate their wellbeing at that time. After a 30-minute break, we asked participants to complete the same items but respond based on their current wellbeing (post-test) (Fig 1, step 11). We repeated data collection a month later, with five of the eight participants asking about their current wellbeing on that day.

### Psychometric analyses

Analyses were iterative and examined item performance, dimensionality, response-category functioning and reliability. Exploratory factor analysis of the 53- and 38-item versions used maximum likelihood with oblique rotations. Rasch analysis assessed item fit, residuals and threshold ordering. Items were removed based on weak item-total correlations, poor Rasch fit, disordered thresholds, low loadings, cross-loadings, redundancy and conceptual coverage. Internal consistency was assessed using Cronbach’s alpha and intraclass correlation coefficients, interpreted alongside content validity and response-process evidence.

### Ethics and dissemination

This study was approved by the national ethics committee, the Medical Research Council of Zimbabwe (#A/2860). We obtained written informed consent from participants aged ≥18 years and assent, plus parental consent, from those <18 years. We removed names and other personal identifiers from transcripts and other data sources prior to analyses. Participants received US$5 bus fare reimbursements.

## Results

### Review findings

The scoping and systematic review has been published elsewhere [17]. We identified 15 positive psychological constructs that had been used to assess ALHIV: body appreciation, confidence, coping, flourishing, meaningfulness, personal control, positive outlook, resilience, self-management, self-compassion, self-concept, self-efficacy, self-esteem, self-worth and transcendence [17]. Resilience, self-esteem and self-efficacy were the most measured constructs [17]. Although the scales had been used in Africa, all but one were developed in high-income settings and were not properly translated and validated before use in Africa.

### Content validity findings

Of 263 candidate items, 217 with complete ratings from all three reviewers were included; 45 duplicate items and one item with a non-numeric rating were excluded. The overall S-CVI/Ave was 0.730, and S-CVI/UA was 0.369. Eighty items (36.9%) achieved unanimous agreement (I-CVI = 1.00), 100 (46.1%) had an I-CVI of 0.67 and were flagged for revision, and 37 (17.1%) had an I-CVI ≤ 0.33, and were considered for removal. Construct-level S-CVI/Ave values were highest for gratitude and perseverance (0.889 each) and lowest for flourishing (0.587) (S2 File).

### Qualitative findings

The initial qualitative piece, which complemented the scoping and systematic review, identified six dominant constructs emerging from the data: connectedness, happiness, hope, motivation, optimism, and perseverance. That initial piece and subsequent ones unravelled insights on the constructs’ context and nuances. For example, young people mentioned that their happiness was defined by life events and was largely “happiness with some tears” given the need to navigate daily struggles such as stigma and discrimination. These contextual factors also threatened another construct, “perseverance”.

An additional insight was that within this setting, “self-acceptance” emerges in relation to others, and is focused on accepting oneself through participating in a community, i.e., (I accept myself within this community). Further, young people mentioned that they considered their own wellbeing in comparison to others in the community (i.e., I am doing well compared to others). As one participant remarked, *‘Yes, I am positive [HIV] but I have a bright future. I can even compare myself with others… I can say that I am not the young person with the worst challenges…”* (Participant #2 ZCS workshop 2). This has important implications for measurement as wellbeing is not only relational but also evaluated in comparison to other possibilities, i.e., (It could be worse, as I see it is for others, therefore for me it is not too bad - even though I am struggling with my wellbeing).

In all discussions, young people highlighted the fluctuating nature of all constructs and cautioned against assuming their continuous presence. They explained this in part because of the precarious contextual conditions which continuously threaten their wellbeing. Finally, discussions highlighted how language around mental health and wellbeing is constrained. For example, it was difficult to differentiate between some constructs (e.g., hope and optimism) or at the very least, to find near-equivalent indigenous terms. These factors have implications for measuring the Zvandiri Character Strength and wellbeing more generally.

### Scale administration findings

Data analysis of the first 530 participants (who completed the initial 53-item scale) showed that responses were highly positive across all items. For each item, the proportion reporting ‘strongly agree’ ranged from 34% to 67%, and the proportion reporting ‘agree’ or ‘strongly agree’ ranged from 81%-98% (all items were positive). A scree plot identified 4 factors, but with low inter-item correlation and multiple items cross-loading across factors. Rasch analysis of polytomous responses showed that the response thresholds were disordered (See S3 File for detailed analyses, including item-level response distributions, factor loadings, Rasch diagnostics, threshold maps and item-retention decisions).

Since the ordinal responses yielded disordered thresholds, we used binary responses (agree/disagree) for the ZCS-38 scale after consulting with young people. 39 young people completed the ZCS-38 (reduced from 53 items). We identified order-related ceiling effects. For the first 15 items on the scale, agreement ranged from 80% to 90%; for items 16-38, agreement ranged from 92% to 100%. This indicated questionnaire fatigue and unreliable responses.

As stated above, a group discussion was held with young people to investigate the causes of ceiling effects. The young people told us the statements were all positive affirmations that encouraged agreement, and that, in the presence of an interviewer, they were concerned about being judged if they did not report positive responses. We added a preamble explaining that there was no need to give responses which pleased the interviewer. This helped us interpret high scores as potentially reflecting a mixture of genuine positive appraisal, encouragement from the item wording, social desirability, demand characteristics and concern about how disagreement would be judged (36, 37). To improve response variability, we reverted to a 4-item Likert scale of responses, using ‘never’, ‘seldom’, ‘some of the time’, and ‘most of the time’ rather than strength of agreement. We benchmarked the responses to ‘in the past two weeks’. To minimise questionnaire fatigue, we reduced the number of items to 25 (ZCS 25).

As stated earlier, eight young people completed a retrospective pretest (Phase 1), and a post-test (Phase 2) of the ZCS 25 scale with the new response options, and five of them completed the ZCS 25 scale again a month later (Phase 3). Scores were calculated as never=0, seldom=1, some of the time=2, most of the time=3 (range 0-75). Retrospective pretest and post-test responses fluctuated, making it difficult to interpret the results (Fig 2). We had expected the Phase 1 score to be lower than that of Phase 2 and Phase 2 to be similar to Phase 3, but there was no evidence of this. Two participants reported their wellbeing after the intervention was considerably lower than it had been retrospectively, but when discussing this qualitatively, they said that, on the contrary, the intervention had positively impacted their wellbeing. We understood this discordance to reflect a challenge with the scale. Ceiling effects continued; four participants reported scores of 70-75.

Taken together, the persistent ceiling effects, fluctuating retrospective ratings, and discordance between qualitative accounts and ZCS 25 scores did not support the use of the ZCS 25 as an outcome measure without substantial further development. We therefore did not proceed to larger-scale quantitative validation at this stage.

**Fig 2:**
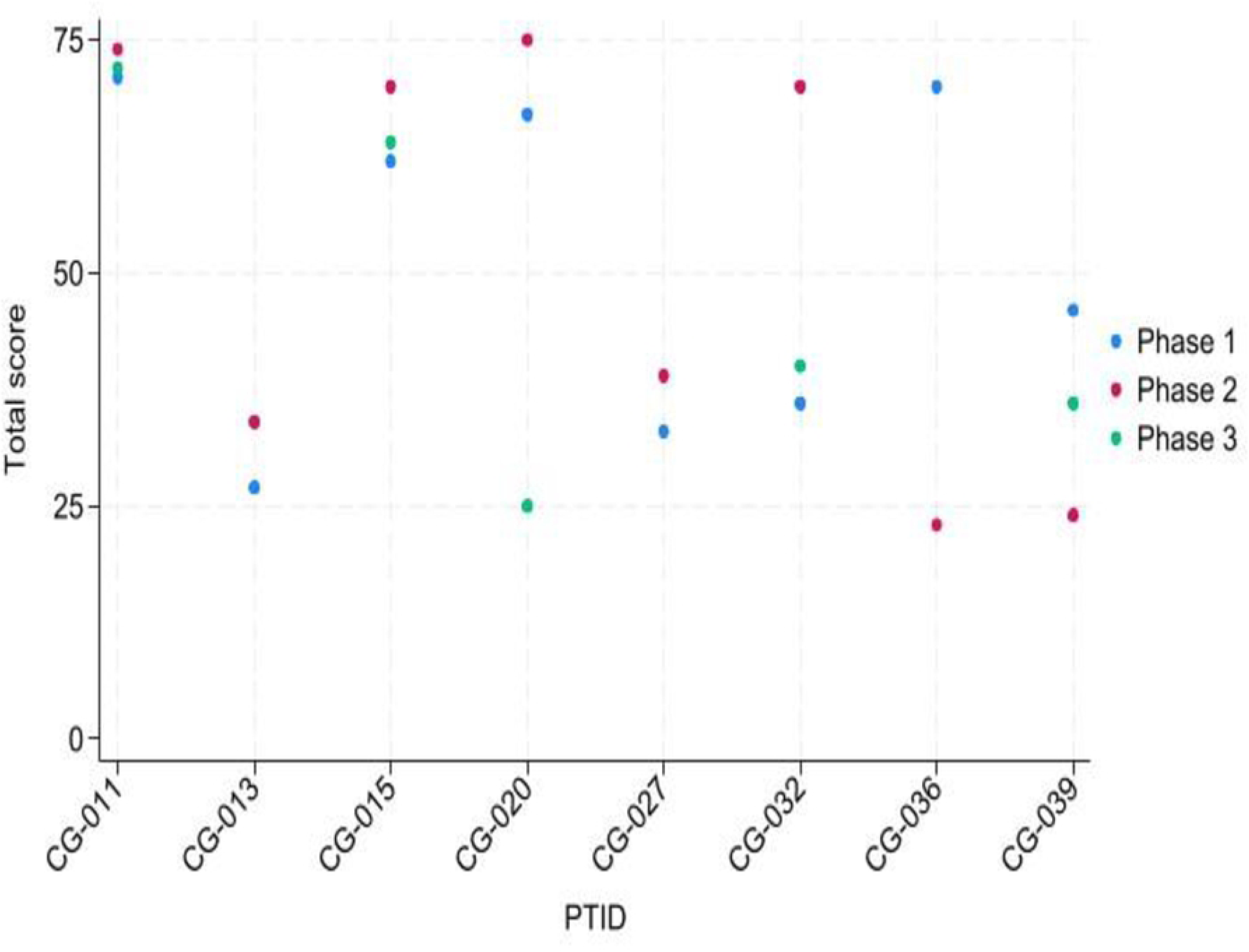
ZCS scale scores for three phases.

## Discussion

We aimed to develop a contextually relevant positive psychological outcome measure for adolescents living with HIV (ALHIV) in Zimbabwe, with potential application to Eastern and Southern Africa more broadly. We followed recognised best-practice standards for patient-reported outcome measure development and validation, including a systematic review and some qualitative interviews to generate candidate items [26]. However, the measure did not achieve the reliability and validity required for confident use. Here, we share lessons learned and recommendations for future studies.

One possible explanation lies in the alignment among the construct definition, item content, response process, and intended score interpretation. We initially explored whether the ZCS could support a single score for positive wellbeing or “character strength”, but the development process brought together several conceptually adjacent yet non-interchangeable domains. In the qualitative work with ALHIV, six dominant constructs emerged: connectedness, happiness, hope, motivation, optimism and perseverance. These domains span social experience, affective state, future orientation, motivational capacity and coping, and therefore may not function as indicators of a single homogeneous latent trait.

The lack of a coherent factor and Rasch structure should not be interpreted as evidence that wellbeing among ALHIV cannot be measured, nor that reflective measurement is inherently inappropriate. Rather, it suggests that the proposed total score was not supported by the available evidence. This may reflect restricted variance due to ceiling effects, disordered response categories, interviewer/social desirability effects and insufficiently specified score interpretation [27, 33–36]. Future work should therefore return to construct pruning, response-process validation and domain-level testing before attempting to justify a single ZCS total score.

The second issue is the construct jangle problem. The broader wellbeing and positive psychology literature includes many overlapping labels: self-worth, self-esteem, self-concept, confidence, self-efficacy, resilience, coping, hope, optimism, perseverance and flourishing. These are not identical constructs, but they are often treated as if their boundaries are self-evident. The jingle-jangle problem is well established in psychological measurement: different labels may be used for highly similar constructs, while similar labels may conceal different constructs [36, 37]. More recent critiques of psychological measurement also warn that weak construct specification and proliferation of partially overlapping measures can undermine validity before statistical modelling even begins [27]. Constructs identified through the systematic review should not automatically be treated as a coherent conceptual framework; rather, they may reflect underlying conceptual heterogeneity. Careful definition, differentiation, and pruning grounded in the lived experiences of ALHIV are therefore required before item generation and structural testing to produce measures that are both conceptually coherent and contextually fit.

Third, the factor and Rasch analyses results may have been compromised by response distributions. If most respondents cluster at the positive end of most items, the correlation matrix may not contain sufficient variation to recover structure, even if meaningful domains exist. Fourth, is the issue of response-process validity. Young people’s comments that the items sounded like positive affirmations, and that they worried about being judged by an interviewer, are response-process evidence. In this case, observed responses may have been shaped by social desirability, demand characteristics, acquiescence, interviewer effects, politeness norms, or loyalty to a valued programme, as well as by underlying wellbeing. Classic work on social desirability and demand characteristics is directly relevant here [38, 39], as is the wider survey-methods literature on how respondents interpret questions and construct answers in context [40–43].

Fifth, cross-cultural survey research shows that response styles, including acquiescence and extreme responding, can vary across cultural and survey contexts [34, 44, 45]. There is also directly relevant evidence from adolescent and HIV-related research in Eastern and Southern Africa that the mode of administration and privacy can shape self-reports [46–48]. Although these studies concern sexual behaviour rather than wellbeing, they are relevant because they demonstrate that adolescent self-report in HIV-related and sexual-health research is vulnerable to administration mode, privacy, perceived judgement and social desirability.

Another key issue is the centrality of relationality. In Africa, the philosophy of *Ubuntu* (humanity) positions personhood as fundamentally constituted through relationships with others [49, 50]. *Ubuntu* therefore frames human flourishing not as an individual possession, but as something realised through mutual recognition, solidarity, compassion, responsibility and harmonious coexistence. From this perspective, wellbeing is not reducible to an internal psychological state located solely within the individual. Instead, it is relational, moral and socially embedded, and, as stated earlier, this has implications for measurement.

Our findings highlight a tension between capturing wellbeing as lived and measuring intervention-relevant change. Qualitative and participatory activities suggested that adolescents experience wellbeing as uneven, contextually-situated and often simultaneous with distress; earlier Zvandiri work similarly found that young people represented feelings as crooked rather than linear trajectories [51, 52], reflecting lives in which adversity continued but support could provide comfort, connection, respite and manageability. Further, adolescence is marked by rapid physical, hormonal, neural, psychosocial, cognitive and emotional transitions [53]. Future ZCS work should therefore clarify whether the intended measure is capturing average wellbeing over a defined period, variability across contexts, or change in specific intervention mechanisms, and should consider context-specific items, repeated assessment, visual trajectory methods or mixed-method interpretation where fluctuation and appraisal are central to the construct.

To interpret our experiences of ZCS scale development, we draw on conceptualisations of wellbeing as multidimensional, relational and dynamic [22, 54–56]. The contribution of the ZCS process is more specific: it shows how these established issues became practically consequential when trying to develop an outcome measure for a relational intervention with ALHIV in Zimbabwe. Future work should therefore move beyond simple adaptation of imported, individual-level measures, but not because quantitative measurement is inappropriate. Rather, the measurement approach needs to be aligned with the intervention theory of change and the intended score interpretation. If the aim is to capture wellbeing as lived, then methods may need to attend to fluctuation, context, relationships and narrative meaning, using qualitative, vignette-based, visual, diary or ecological momentary assessment (EMA)-informed approaches where appropriate [57–59]. If the aim is to evaluate intervention benefit, however, the measure must also be capable of detecting theorised change in specific domains that an intervention seeks to influence, such as connectedness, self-worth, hope, treatment confidence, relational safety or manageability. These aims are related but not identical. The practical lesson is that future ZCS measurement must decide what kind of change it is trying to detect, at what level, over what time frame, and using which combination of quantitative and qualitative methods.

Importantly, our results should not be read as evidence that positive wellbeing cannot be measured among ALHIV in Zimbabwe, nor that rigorous psychometric methods are irrelevant. We already have good examples to show that local concepts of self, family, spirituality, belonging and wellbeing can be operationalised psychometrically, but only when construct development precedes scoring. These include for example, the Shona Symptom Questionnaire in Zimbabwe, which developed an indigenous measure of common mental disorders from local idioms of distress [60]; the Hua Oranga, a Maori mental health outcome measure grounded in Maori models of health [61] and the mixed-methods youth mental health scale development in Rwanda [62]. In that sense, the main contribution of this study is methodological as much as it is substantive.

### Strengths and limitations

A strength of our approach is that we employed recognised best-practice standards for patient-reported outcome measure development and validation, including a systematic review and allied qualitative and participatory components to generate candidate items. A potential limitation is the small number of young people who took part in the three phases of ZCS 25 completion, necessitated by the desire to conduct an in-depth exploration of ceiling effects. Also, we did not compare various modes of scale administration. Future initiatives could include fuller item- and domain-level documentation, participant-characteristic reporting, administration-mode testing, domain-specific analyses and systematic response-process evidence.

### Conclusion

We implemented a series of initiatives to collaboratively develop a youth-focused, locally relevant, quantitative instrument to evaluate the Zvandiri Character Strength. Our experiences, consistent with those from similar settings [19, 63], underscore the necessity of appropriately tailoring (wellbeing) scales originally designed elsewhere prior to their application to this context. Future ZCS measurement initiatives should, therefore, move beyond the simple adaptation of imported wellbeing scales and begin with a clear definition of each construct, locally grounded language, response-process evidence, and an explicit decision about what the measure is intended to capture: lived wellbeing, domain-specific positive outcomes, or intervention-relevant change. This decision should guide item content, recall period, response options, administration mode, scoring, and the balance between quantitative and qualitative evidence.

## Data Availability

All data related to tool development have been submitted as supporting files.

## Acknowledgements

The authors would like to thank the young people who took part in the various activities. Thank you also to everyone at Zvandiri who supported and facilitated this study.

## Supporting information

S1_File: 53-item scale spanning 14 constructs

S2_File: Panel of experts’ content validation results

S3_File: Detailed psychometric evaluation results

